# Real-world insights into antiobesity medications in an online patient community: a feasibility study

**DOI:** 10.64898/2026.01.26.26344623

**Authors:** Bethany Kalich, Marisa Cruz, Stanley Shaw, Jeanine M. Cordova, Hengyi Ke, Michael Mina, Brian Chen, Paul Burton

## Abstract

Decentralized clinical trials offer a scalable approach to evaluate patient research in real-world settings. The COSMOS–DIGITAL study (NCT06761703) was designed to assess the feasibility of recruiting patients using antiobesity medications from an online patient community and aimed to evaluate participants’ willingness to consent to and complete fully decentralized surveys and at-home self-blood testing. Three-quarters (n = 151; 75.5%) of participants completed all self-collected capillary blood samples, Patient-Reported Outcomes in Obesity (PROS) surveys, and nausea-related surveys. Over 83% of participants completed the surveys (PROS: n = 167; 83.5%; daily nausea: n = 168; 84.0%), and 93% (n = 142/152) of collected blood samples were sufficient or partially sufficient for testing. Overall, participants were satisfied with the at-home blood collection device. Digitally enabled, fully decentralized studies can capture blood samples and survey responses as well as important aspects of patient treatment experiences, adding value to data collected in randomized controlled trials.

---

Digitally enabled, decentralized clinical trials (DCTs) perform some or all study procedures at a location other than a traditional clinical trial site.^1-3^ Many clinical trials adopted DCT approaches during the coronavirus disease 2019 (COVID-19) pandemic, including conducting all procedures in the participants’ home (fully DCT).^4-6^ However, limited evidence outside the COVID-19 pandemic exists about patient willingness to participate in fully decentralized screening, consent, and other study procedures (including blood sampling).

Despite broad uptake of antiobesity medications (AOMs), data about their real-world use remains limited. To better understand the day-to-day patient experience on AOMs, we performed a fully decentralized study in individuals taking a GLP-1 RA–based therapy (semaglutide or tirzepatide) for obesity or overweight (Community Outreach Support and engageMent using Online Strategies - Decentralized InsiGhts Into Therapy Adoption and vaLue [COSMOS–DIGITAL]; NCT06761703). Approximately 276,000 emails were sent by the time enrollment was closed, 474 individuals completed the online screening, and 200 participants were recruited in 8 days (Figure 1). On day 1 of the study, an at-home self-obtained capillary blood sample device (Touch Activated Phlebotomy [TAP^®^]), a Patient-Reported Outcomes in Obesity (PROS) survey, a patient activation (Partners in Health [PiH]) survey, and a nausea questionnaire were sent to each participant. On day 30, the same surveys as well as an additional survey on patient satisfaction with the TAP^®^ device were sent to each participant. Participants were also sent a nausea diary with questions to be answered daily (Figure 2).

**Fig. 1.**
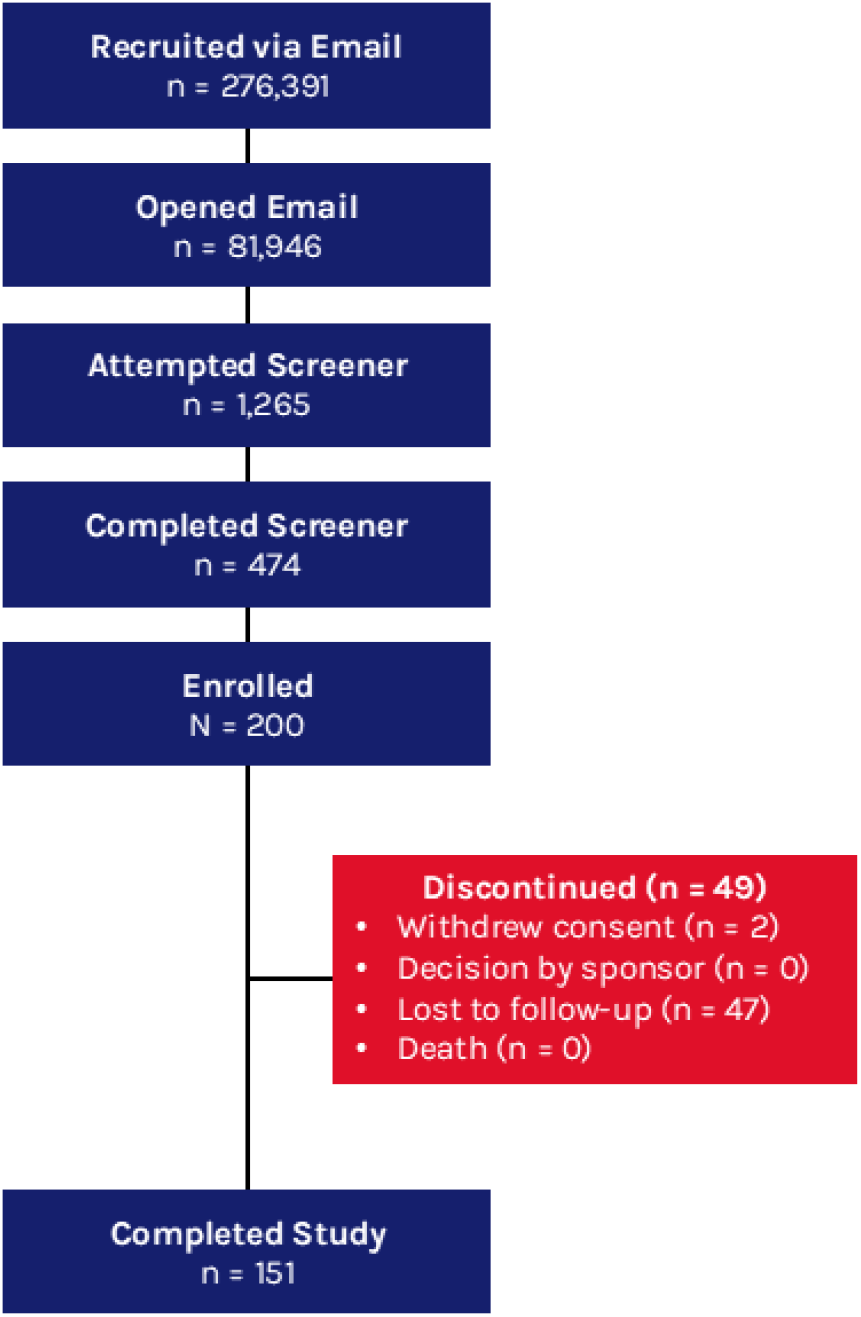
Consort Diagram

**Fig. 2.**
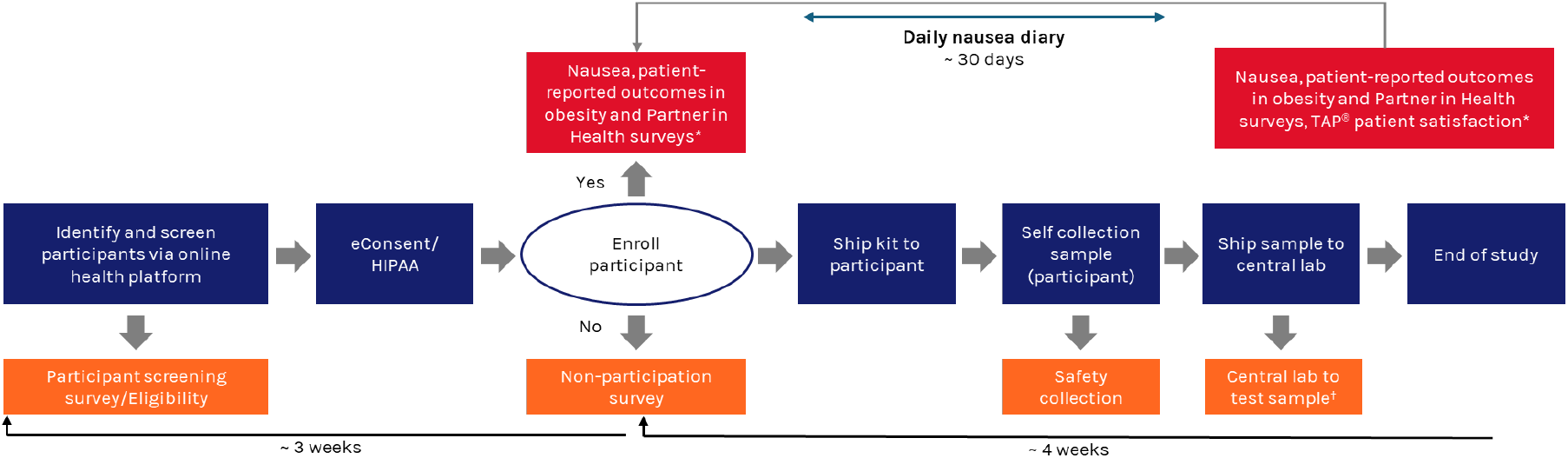
Study Design. *Surveys completed within approximately 7 days of receipt. ^†^Total cholesterol, HDL-C, LDL-C, triglycerides, HbA_1c_, creatinine, and hs-CRP. HbA_1c_, hemoglobin A_1c_; HDL-C, high-density lipoprotein cholesterol; LDL-C, low-density lipoprotein cholesterol; HIPAA, Health Insurance Portability and Accountability Act; hs-CRP, high-sensitivity C-reactive protein; TAP^**®**^, Touch Activated Phlebotomy.

The cohort was predominantly female (n = 172; 86.0%), had a mean (standard deviation [SD]) age of 54 (10.8) years, and included 8.0% (n = 16) Black and 5.5% (n = 11) Latino/Hispanic participants. The mean (SD) body mass index (BMI) was 33.9 (8.1) kg/m^2^ (64.5% [n = 129] reported a BMI > 30 kg/m^2^). Participants were evenly split between taking semaglutide or tirzepatide (n = 102 [51.0%] vs n = 98 [49.0%]). Most participants reported using the medications for weight loss alone (n = 139 [69.5%]) vs for both weight loss and diabetes management (n = 61 [30.5%). At the time of enrollment, 41.0% (n = 82) of participants had been on an AOM for more than 6 months. In the day 1 and 30 surveys, participants self-reported a high prevalence of cardiometabolic comorbidities, including hypertension (n = 86 [43.0%]), hyperlipidemia (n = 86 [43.0%]), and diabetes (n = 61 [30.5]%; Table 1).

**Table 1.** Baseline Demographics.

|  | **Overall**<br>**N = 200** |
| --- | --- |
| **Age, mean (SD)** | 54 (10.8) |
| **Sex** |  |
| Female | 172 (86.0) |
| Male | 28 (14.0) |
| **Race** |  |
| American Indian or Alaskan Native | 3 (1.5) |
| Asian or Asian American | 4 (2.0) |
| Black or African American (non-Hispanic) | 16 (8.0) |
| Latino or Hispanic | 11 (5.5) |
| Native Hawaiian or Other Pacific Islander | 0 (0.0) |
| White | 175 (87.5) |
| Other* | 0 (0.0) |
| Unknown | 1 (0.5) |
| **BMI category (kg/m<sup>2</sup>)<sup>†</sup>** |  |
| Under/healthy weight (< 25) | 19 (9.5) |
| Overweight (≥ 25 to < 30) | 52 (26.0) |
| Obesity, class I (≥ 30 to < 35) | 56 (28.0) |
| Obesity, class II (≥ 35 to < 40) | 39 (19.5) |
| Obesity, class III (≥ 40) | 34 (17.0) |
| **Employment** |  |
| Full-time employment | 109 (54.5) |
| Part-time employment | 24 (12.0) |
| Not employed | 67 (33.5) |
| **GLP-1 RA–based therapy** |  |
| Tirzepatide | 98 (49.0) |
| Semaglutide | 102 (51.0) |
| **Purpose of AOM** |  |
| Weight management | 139 (69.5) |
| Weight management and diabetes | 61 (30.5) |
| **Duration of AOM** |  |
| Less than 1 month | 20 (10.0) |
| 1 to 3 months | 52 (26.0) |
| 3 to 6 months | 46 (23.0) |
| More than 6 months | 82 (41.0) |
| **Medical history<sup>‡</sup>** |  |
| Overweight (BMI > 25 kg/m<sup>2</sup> to < 30 kg/m<sup>2</sup>) | 71 (35.5) |
| Obesity (BMI > 30 kg/m<sup>2</sup>) | 152 (76.0) |
| High blood pressure | 86 (43.0) |
| High cholesterol | 86 (43.0) |
| Diabetes | 61 (30.5) |

Data are n (%) unless otherwise indicated.
\*“Other” indicates Mediterranean (not European) or Southeast Asian.
<sup>†</sup>BMI calculated at the time of analysis based on height and weight submissions.
<sup>‡</sup>Patient-reported.
AOM, antiobesity medication; BMI, body mass index; N, number of participants in the analysis set; n, number of participants with observed data; SD, standard deviation.

Of the 200 enrolled participants, three-quarters (n = 151 [75.5%]) completed all study procedures, including surveys and returning the self-administered capillary blood sample (Figure 1). All participants (n = 200 [100%]) provided a complete medical history on days 1 and 30, and over 83% of participants completed the day 1 and day 30 PROS (n = 167 [83.5%]) and PiH (n = 166; 83.0%) surveys as well as the day 30 TAP^®^ satisfaction survey (n = 167 [83.5%]) and daily nausea diaries (n = 168 [84.0%]). Fewer responses were received for the nausea questionnaire (n = 96 [48.0%]; Table 2).

**Table 2.** Percentage of Participants Who Completed Survey Components.

|  | **Overall**<br>**N = 200** |
| --- | --- |
| **Medical history** |  |
| Yes | 200 (100) |
| No | 0 (0.0) |
| **PROS questionnaire** |  |
| Yes | 167 (83.5) |
| No | 33 (16.5) |
| **PiH questionnaire** |  |
| Yes | 166 (83.0) |
| No | 34 (17.0) |
| **TAP® satisfaction survey** |  |
| Yes | 167 (83.5) |
| No | 33 (16.5) |
| **Nausea and nausea management\*** |  |
| Yes | 96 (48.0) |
| No | 104 (52.0) |
| **Daily nausea diary** |  |
| Yes | 168 (84.0) |
| No | 32 (16.0) |

Data are n (%).
“Yes” indicates that the participant completed all questions within the respective survey component on all required survey days, and “No” indicates incomplete or missing responses for any question in the survey component on any of the required survey days.
\*Completion rule for nausea and nausea management: if a participant’s response was “1 = no nausea” in question 9 in one survey, then this participant was automatically treated as having completed all questions for nausea management in that survey. If a participant selected some degree of nausea (ranging from 2 to 10) in one survey, and they responded to all follow-up questions related to nausea management, then this section was considered complete for that survey. To report “Yes” in the table, the participant needed to complete the nausea and nausea management section in both the day 1 and day 30 surveys.
N, number of participants in the analysis set; n, number of participants with observed data; PiH, Partners in Health; PROS, patient-reported outcomes in obesity; TAP®, Touch Activated Phlebotomy.

With written and video instructions but no additional supervision or proctoring, 76.0% (n = 152) of participants submitted an at-home, self-collected capillary blood sample. Of these, 93.4% (n = 142) of the samples had sufficient quantity for full or partial testing, with 61.2% (n = 93) of the samples usable for full laboratory analysis (hemoglobin A_1c_, total cholesterol, low-density lipoprotein cholesterol, high-density lipoprotein cholesterol, triglycerides, creatinine, and high-sensitivity C-reactive protein). A majority of participants (n = 133 [79.6%]) agreed or strongly agreed that they would be willing to self-obtain a blood sample monthly or quarterly if required for a study, and 69.5% (n = 116) agreed or strongly agreed that the self-administered blood draw was preferable to phlebotomy at a trial site (Table 3). Results from submitted samples were within conventional ranges and consistent with expectations for the study population (Supplemental Table 1).^7^

**Table 3.** Patient Satisfaction With the TAP^®^ Device for Self-obtained Capillary Blood Sample, of Participants Who Used the TAP^®^ Device.

|  | **Day 30**<br>**N = 167** |
| --- | --- |
| **Participants would use the home-based self-collection method again in the future, n (%)** |  |
| Strongly agree | 87 (52.1) |
| Agree | 48 (28.7) |
| Neutral | 18 (10.8) |
| Disagree | 6 (3.6) |
| Strongly disagree | 8 (4.8) |
| **Participants prefer the at-home TAP<sup>®</sup> device over the clinic blood draw, n (%)** |  |
| Strongly agree | 72 (43.1) |
| Agree | 44 (26.3) |
| Neutral | 29 (17.4) |
| Disagree | 14 (8.4) |
| Strongly disagree | 8 (4.8) |
| **Participants feel confident that they collected the TAP<sup>®</sup> sample correctly, n (%)** |  |
| Strongly agree | 79 (47.3) |
| Agree | 51 (30.5) |
| Neutral | 18 (10.8) |
| Disagree | 12 (7.2) |
| Strongly disagree | 7 (4.2) |
| **Participants would use the home collection method monthly or quarterly if needed, n (%)** |  |
| Strongly agree | 79 (47.3) |
| Agree | 54 (32.3) |
| Neutral | 20 (12.0) |
| Disagree | 9 (5.4) |
| Strongly disagree | 5 (3.0) |
| **Participants would use the home-based blood test if available commercially, n (%)** |  |
| Strongly agree | 71 (42.5) |
| Agree | 51 (30.5) |
| Neutral | 29 (17.4) |
| Disagree | 11 (6.6) |
| Strongly disagree | 5 (3.0) |
| **Participants recommend the TAP<sup>®</sup> device on a scale of 0–10** |  |
| Mean (SD) | 7.5 (2.7) |
| Median (Q1, Q3) | 8.0 (5.0, 10.0) |

Data are n (%) unless otherwise indicated.
N, number of participants in the analysis set; n, number of participants with observed data; Q1, first quartile; Q3, third quartile; SD, standard deviation; TAP<sup>®</sup>, Touch Activated Phlebotomy.

For participant PROS, the most common response for most measures was “mildly bothered” (including common physical activities, bodily pain, sleep, and social interactions). Notable exceptions were self-esteem (“considerably bothered” was most common) as well as sexual life and discrimination or discourteous behavior (“not bothered” was most common for both; Table 4). Participants generally scored high in measures of patient engagement and activation (Supplemental Table 2).

**Table 4.** Patient-Reported Outcomes in Obesity Survey Results.

|  | **Day 1**<br>**N = 189** | **Day 30**<br>**N = 167** |
| --- | --- | --- |
| **Common physical activities** |  |  |
| Considerably bothered | 39 (20.6) | 29 (17.4) |
| Moderately bothered | 50 (26.5) | 38 (22.8) |
| Mildly bothered | 72 (38.1) | 62 (37.1) |
| Not bothered | 28 (14.8) | 38 (22.8) |
| **Bodily pain** |  |  |
| Considerably bothered | 39 (20.6) | 33 (19.8) |
| Moderately bothered | 55 (29.1) | 40 (24.0) |
| Mildly bothered | 61 (32.3) | 59 (35.3) |
| Not bothered | 34 (18.0) | 35 (21.0) |
| **Discrimination or discourteous behavior** |  |  |
| Considerably bothered | 18 (9.5) | 18 (10.8) |
| Moderately bothered | 36 (19.0) | 25 (15.0) |
| Mildly bothered | 49 (25.9) | 53 (31.7) |
| Not bothered | 86 (45.5) | 71 (42.5) |
| **Sleep** |  |  |
| Considerably bothered | 25 (13.2) | 14 (8.4) |
| Moderately bothered | 40 (21.2) | 35 (21.0) |
| Mildly bothered | 72 (38.1) | 67 (40.1) |
| Not bothered | 52 (27.5) | 51 (30.5) |
| **Sexual life** |  |  |
| Considerably bothered | 37 (19.6) | 30 (18.0) |
| Moderately bothered | 54 (28.6) | 39 (23.4) |
| Mildly bothered | 42 (22.2) | 42 (25.1) |
| Not bothered | 56 (29.6) | 56 (33.5) |
| **Normal social interaction** |  |  |
| Considerably bothered | 15 (7.9) | 9 (5.4) |
| Moderately bothered | 39 (20.6) | 36 (21.6) |
| Mildly bothered | 68 (36.0) | 67 (40.1) |
| Not bothered | 67 (35.4) | 55 (32.9) |
| **Work, school, or other daily activities** |  |  |
| Considerably bothered | 18 (9.5) | 9 (5.4) |
| Moderately bothered | 37 (19.6) | 32 (19.2) |
| Mildly bothered | 68 (36.0) | 56 (33.5) |
| Not bothered | 66 (34.9) | 70 (41.9) |
| **Self-esteem** |  |  |
| Considerably bothered | 75 (39.7) | 54 (32.3) |
| Moderately bothered | 50 (26.5) | 50 (29.9) |
| Mildly bothered | 53 (28.0) | 42 (25.1) |
| Not bothered | 11 (5.8) | 21 (12.6) |

Data are n (%).
N, number of participants in the analysis set; n, number of participants with observed data.

Participants reported in the day 1 and day 30 surveys that nausea severity (on a 10-point scale; 10 being the worst) was relatively mild (mean [SD] of 2.9 [2.0] and 2.8 [1.7] on days 1 and 30, respectively). Notably, many participants reported significant use of anti-nausea remedies (n = 48 [25.4%] and n = 61 [36.5%] on days 1 and 30, respectively), including prescription medications (n = 28 [14.8%] and n = 33 [19.8%] on days 1 and 30, respectively; Supplemental Table 3), and this trend was numerically higher during months where titration of therapy was likely (Supplemental Figure 1a). When asked to estimate how many days in the past month they experienced nausea, 29.3% (n = 49) reported 0 to 5 days, and 42.5% (n = 71) reported 6 or more days per month (19.2% [n = 32], 14.4% [n = 24], 6.0% [n = 10], and 3.0% [n = 5] reported nausea lasting 6–10 days, 11–15 days, 16–20 days, and 21+ days, respectively; day 30 survey). Participants reported that nausea symptoms persisted regardless of AOM treatment duration (1 to > 6 months; Supplemental Table 3 and Supplemental Figure 1b).

Participants also completed an electronic daily diary of their nausea symptoms. Similar to the above results, diary entries indicated that nausea symptom severity, impact of nausea on quality of life, and requirement for anti-nausea medications persisted at comparable levels regardless of AOM treatment duration (Supplemental Figure 2).

Together, these results highlight the feasibility of digitally enabled DCT approaches to engage participants in a variety of study procedures, including granting e-consent, obtaining self-administered capillary blood samples, completing surveys, and providing information about important medication-related adverse events. This study used a deliberately minimal support model featuring only asynchronous communication (such as emails) and written and video instructions (for self-blood collection); the degree of engagement observed here may help establish a lower bound of engagement rates and inform the design of DCTs following the COVID-19 pandemic. Synchronous communications, tele-visits, live proctoring (eg, for self-obtained blood sampling or other procedures), and other digital tools would be expected to improve upon this baseline. Participant enthusiasm for self-obtained blood sampling was high and supports the promise of this capability to enable biospecimen collection as part of patient-focused, at-home decentralized trials.

Previous real-world evidence approaches to assessing side effects associated with GLP-1 RA–based therapy have used electronic health records and insurance claims, the US Food and Drug Administration Adverse Event Reporting System, and the National Institutes of Health “All of Us” database.^8-10^ While valuable for identifying broad usage patterns or safety signals, these data analyses provide limited views into patient experience, potentially because patient-coping strategies for medication side effects may not be captured in electronic health record or administrative databases. In contrast, the current study leverages patient PROS and responses through digital engagement to offer unique insights into the patient experience regarding side effects from GLP-1 RA–based therapy use for obesity or overweight.

Limitations of this study include reliance on self-reported data for AOM administration and nausea-related adverse events and recruitment in the US only. Furthermore, recruitment from a generalized online patient community could have introduced bias toward participants who are comfortable with online surveys and naturally more involved in their healthcare. Despite these limitations, online recruitment was rapid, resulted in a representative, multi-ethnic study population with 8.0% Black or African American participants (compared to ∼5% and ∼8% in the global pivotal semaglutide and tirzepatide phase 3 studies, respectively), and exhibited a high prevalence of major cardiometabolic comorbidities of obesity.^11-13^

This study illustrates the potential of digitally enabled, decentralized research to understand the real-world patient journey on therapies such as AOMs. Insights from this digitally enabled DCT can inform future clinical trials and practice as well as shared decision-making and support strategies to improve the experience for individuals initiating or continuing certain therapies.

## Methods

### Study Design

This was an unblinded, decentralized, digitally led, nonrandomized study (ClinicalTrials.gov ID: NCT06761703; registered January 7, 2025). All individuals who consented to participate in the study were sent a self-collection blood draw device (TAP^®^ device) from YourBio Health (Medford, MA) and completed surveys at baseline and 30 days via the Inspire digital health platform (inspire.com; Arlington, VA). Inspire provides online educational information and peer support for hundreds of condition-specific patient communities. Participants also completed a daily diary reporting nausea and vomiting symptoms and management. The initial screening survey confirmed the use of a GLP-1 RA–based therapy for obesity or overweight and eligibility criteria. The end of the study was defined as the date when the last participant completed the last scheduled activity. The reporting period spanned from November 14, 2024, through January 6, 2025 (Figure 2).

### Patient Consent

The study protocol, protocol amendments, informed consent form, and other relevant documents were reviewed and approved by Advarra (Columbia, MD, USA). Due to the decentralized nature of this study, electronic consent (21 CFR Part 11 compliant) was used. All participants were required to electronically sign and date an institutional review board–approved informed consent form before any study-specific procedures were performed.

### Key Inclusion and Exclusion Criteria

Eligible participants had to be 18 to 80 years of age, live in the US, and consent to participate in the COSMOS–DIGITAL study through the Inspire platform. Participants also self-reported the use of semaglutide (Wegovy^®^) or tirzepatide (Zepbound^®^) for the indication of weight management/obesity and reported their intention to continue the therapy for at least 30 days. Participants with diabetes taking semaglutide or tirzepatide for obesity or overweight were eligible, but diabetes was not a primary indication in this study.

Key exclusion criteria included current treatment with another investigational device or drug study or participation in a clinical trial; known history of bleeding diathesis or any coagulation disorder; or history of skin disorders, abnormal skin integrity, or atypical skin health within the areas where blood was to be drawn on the upper arm. Additionally, participants with self-reported sensitivity and/or allergy to any of the components of the self-collection blood draw device, including stainless steel or elements commonly found in stainless steel; self-reported fear of blood; or self-reported circulatory conditions causing difficulty drawing capillary blood, were excluded.

### Objectives and Endpoints

The primary objective of the study was to determine the willingness of participants to consent to at-home self-collection blood testing, with the primary endpoint assessing the number of participants who completed the blood draw. The secondary objective was to determine the ability of participants to complete survey components and to satisfactorily complete an at-home blood draw sampling. For this objective, the completion of surveys on days 1 and 30 and sufficiency of blood volume for analysis were assessed.

Exploratory outcomes included descriptive analyses of participant-reported nausea, impact of AOM therapy duration on nausea severity, obesity-related quality of life, and PiH score as a proxy for activation score. Additional exploratory outcomes included anti-nausea medication use and patient satisfaction with the self-collected blood draw device. The survey responses, including responses to questions related to nausea, the PROS survey addressing obesity-related quality of life, the PiH survey addressing patient activation, and questions addressing satisfaction with the at-home self-collection blood draw device, served as exploratory endpoints.

### Patient Information and Surveys

Self-reported demographic data were collected during screening, and medical history and physical measurements (ie, height and weight) were reported by participants on survey days 1 and 30. All surveys were sent to participants electronically through the Inspire platform. Day 1 surveys were sent to participants upon enrollment in the study, and day 30 surveys were sent approximately 30 days later; survey completion was required within 7 days of receipt for both. A survey about nausea symptoms and their effects was to be answered daily.

Five surveys were sent to participants during this study. A 13-point, one-time survey on AOM use and nausea was sent on day 1 (Supplemental Appendix 1). The nausea scale survey consisted of four sections: the nausea score, where the participant rated their nausea (1 to 10; 1 being the best, and 10 being the worst); the negative impact of nausea on life (eg, physical activity, body pain, behavior, sleep, sexual life, social interaction, work/school/other daily activities, and self-esteem); the use of nausea medication or food remedy; and if the participant skipped an AOM dose due to nausea (Supplemental Appendix 2). The PROS survey was an 8-item survey covering common physical activities; bodily pain; discrimination or discourteous behavior; sleep; sexual life; normal social interaction; work, school, or other daily activities; and self-esteem (Supplemental Appendix 3). The PiH Scale survey was a 12-item survey that measured the ability of patients with chronic conditions to self-manage their disease (Supplemental Appendix 4). The TAP^®^ survey consisted of seven questions on how participants felt about using the TAP^®^ device for at-home self-administered blood draws (Supplemental Appendix 5). A participant was considered to have completed the study if they had completed the last scheduled activity applicable to them. Participants were compensated for their time based on completion of study activities.

### TAP^®^ Blood Collection

A TAP^®^ device for blood collection, written instructions, and a QR code linking to a training video on the use of the TAP^®^ device were provided to each participant upon enrollment. Participants were expected to collect their own blood sample within approximately 7 days after receipt of the kit. After collection, the samples were shipped to a certified laboratory (Roche, Basel, Switzerland) for analysis.

### Statistical Analysis

The primary and secondary endpoints were summarized using descriptive statistics such as number and percentage. Participant-reported information, such as demographics, height, weight, nausea score, nausea severity, quality of life, outcomes in the PROS and PiH surveys, and laboratory results from the capillary blood samples, was reported using descriptive statistics. Participant-reported height and weight were used to calculate BMI, which was reported descriptively.

## Data Availability

Qualified researchers may request data from Amgen clinical studies. Complete details are available at https://www.amgen.com/science/clinical-trials/clinical-data-transparency-practices/clinical-trial-data-sharing-request.

## Acknowledgments

The authors would like to thank the participants and the corresponding staff who participated in this study. Writing and editorial assistance were provided by Stacie Meaux, PhD, of Red Nucleus, and were funded by Amgen Inc. This study was funded by Amgen Inc.

## Author Contributions

**BK** contributed to the conception and design of the study and to the analysis and interpretation of the data.

**MC** contributed to the conception and design of the study and to the analysis and interpretation of the data.

**SS** contributed to the conception and design of the study and to the analysis and interpretation of the data.

**JMC** contributed to the conception and design of the study and to the analysis and interpretation of the data.

**HK** contributed to the analysis and interpretation of the data.

**MM** contributed to the conception and design of the study and to the collection of patient data.

**BC** contributed to the analysis and interpretation of the data and to the collection of patient data.

**PB** contributed to the conception and design of the study and to the analysis and interpretation of the data.

## Competing Interests

**BK, SS, JMC, HK, and PB** are employees of Amgen Inc., and own stock.

**MC** is a former employee of Amgen Inc. and owns stock.

**MM** is an employee of YourBio Health, Inc.

**BC** has no competing interests.

## Notes

### Clinical Trial

NCT06761703

### Author Declarations

The IRB of Advarra (Columbia, MD, USA) gave ethical approval for this work.

