## Supplemental Material for "Real-world insights into antiobesity medications in an online patient community: a feasibility study"

**Supplemental Table 1 | Summary of Chemistry Laboratory Values**

| | Overall<br>N = 200 |
|---|---|
| **Cholesterol (mg/dL), n = 114** | |
| Mean (SD) | 180.8 (44.2) |
| Median (Q1, Q3) | 183.5 (152.0, 207.0) |
| **HDL (mg/dL), n = 111** | |
| Mean (SD) | 56.4 (13.1) |
| Median (Q1, Q3) | 56.0 (48.0, 65.0) |
| **Triglycerides (mg/dL), n = 101** | |
| Mean (SD) | 118.2 (57.5) |
| Median (Q1, Q3) | 106.0 (78.0, 140.0) |
| **LDL (mg/dL), n = 100** | |
| Mean (SD) | 97.7 (37.8) |
| Median (Q1, Q3) | 98.0 (72.0, 123.5) |
| **$HbA_{1c}$, n = 139** | |
| Mean (SD) | 5.39 (0.62) |
| Median (Q1, Q3) | 5.20 (5.00, 5.60) |
| **Creatinine (mg/dL), n = 107** | |
| Mean (SD) | 0.936 (0.289) |
| Median (Q1, Q3) | 0.900 (0.780, 1.020) |
| **hs-CRP (mg/dL), n = 97** | |
| Mean (SD) | 0.39 (1.53) |
| Median (Q1, Q3) | 0.10 (0.00, 0.30) |

$HbA_{1c}$, hemoglobin $A_{1c}$; HDL, high-density lipoprotein; hs-CRP, high-sensitivity C-reactive protein; LDL, low-density lipoprotein; Q1, first quartile; Q3, third quartile; SD, standard deviation.

**Supplemental Table 2 | Partners in Health Survey Results for Patient Activation**

| | Day 1<br>N = 189 | Day 30<br>N = 167 |
|---|---|---|
| **Participants' knowledge of their health condition(s)** | | |
| Mean (SD) | 7.1 (1.1) | 7.2 (1.1) |
| Median (Q1, Q3) | 7.0 (6.0, 8.0) | 8.0 (7.0, 8.0) |
| **Participants' knowledge about the treatment and medication for their health condition(s)** | | |
| Mean (SD) | 6.7 (1.2) | 7.0 (1.1) |
| Median (Q1, Q3) | 7.0 (6.0, 8.0) | 7.0 (6.0, 8.0) |
| **Participants follow the medications or treatments prescribed by their doctor** | | |
| Mean (SD) | 7.4 (0.9) | 7.5 (0.9) |
| Median (Q1, Q3) | 8.0 (7.0, 8.0) | 8.0 (7.0, 8.0) |
| **Participants share decisions about their health condition(s) with their doctor** | | |
| Mean (SD) | 7.4 (1.1) | 7.4 (0.9) |
| Median (Q1, Q3) | 8.0 (7.0, 8.0) | 8.0 (7.0, 8.0) |
| **Participants get culturally appropriate services from health professionals** | | |
| Mean (SD) | 7.1 (1.3) | 7.1 (1.4) |
| Median (Q1, Q3) | 8.0 (7.0, 8.0) | 8.0 (7.0, 8.0) |
| **Participants attend appointments as asked by their doctor** | | |
| Mean (SD) | 7.6 (0.9) | 7.7 (0.7) |
| Median (Q1, Q3) | 8.0 (8.0, 8.0) | 8.0 (8.0, 8.0) |
| **Participants keep track of their symptoms and early warning signs** | | |
| Mean (SD) | 6.1 (2.0) | 6.4 (1.7) |
| Median (Q1, Q3) | 6.0 (5.0, 8.0) | 7.0 (6.0, 8.0) |
| **Participants' action on worsening symptoms** | | |
| Mean (SD) | 6.7 (1.5) | 6.8 (1.5) |
| Median (Q1, Q3) | 7.0 (6.0, 8.0) | 7.0 (6.0, 8.0) |
| **Participants manage the effect of their health condition on their physical activity** | | |
| Mean (SD) | 5.8 (1.7) | 6.1 (1.8) |
| Median (Q1, Q3) | 6.0 (5.0, 7.0) | 6.0 (5.0, 8.0) |
| **Participants manage the effect of their health condition on their social life** | | |
| Mean (SD) | 5.5 (2.0) | 5.8 (2.0) |
| Median (Q1, Q3) | 6.0 (4.0, 7.0) | 6.0 (5.0, 7.0) |

Scale of 0 to 8; 0 being very little/never/not very well and 8 being a lot/always/very well.
N, number of participants in the analysis set; SD, standard deviation; Q1, first quartile; Q3, third quartile.

**Supplemental Table 3 | Self-Reported Nausea and Nausea Management**

| | Day 1<br>N = 189 | Day 30<br>N = 167 |
|---|---|---|
| **Nausea scale*** | | |
| Mean (SD) | 2.9 (2.0) | 2.8 (1.7) |
| Median (Q1, Q3) | 2.0 (1.0, 5.0) | 2.0 (1.0, 4.0) |
| **Nausea management, n (%)** | | |
| Took any nausea medication† | 48 (25.4) | 61 (36.5) |
| Took prescription medication | 28 (14.8) | 33 (19.8) |
| Took over-the-counter antihistamine | 16 (8.5) | 10 (6.0) |
| Took over-the-counter stomach relief | 6 (3.2) | 30 (18.0) |
| Used natural food remedies | 33 (17.5) | 56 (33.5) |
| Stopped taking AOM | 0 (0.0) | 0 (0.0) |
| Delayed taking AOM | 4 (2.1) | 10 (6.0) |
| Skipped AOM dose | 2 (1.1) | 3 (1.8) |
| Contacted doctor | 6 (3.2) | 7 (4.2) |
| **Days participants felt nauseous, n (%)** | | |
| 0–5 days | 77 (40.7) | 49 (29.3) |
| 6–10 days | 30 (15.9) | 32 (19.2) |
| 11–15 days | 12 (6.3) | 24 (14.4) |
| 16–20 days | 6 (3.2) | 10 (6.0) |
| 21+ days | 3 (1.6) | 5 (3.0) |
| **Description of nausea, n (%)** | | |
| First few weeks, then went away | 34 (18.0) | 15 (9.0) |
| Every time with medication | 33 (17.5) | 50 (29.9) |
| No nausea | 2 (1.1) | 1 (0.6) |
| Other | 58 (30.7) | 54 (32.3) |

*Rated 1 to 10; 1 being the best and 10 being the worst.

†Sum of participants across "nausea management measure," "took nausea medication," and "description of nausea" subcategories may exceed the total number of participants in the full analysis set, as participants are allowed to select more than one option.

AOM, antiobesity medication; N, number of participants in the analysis set; n, number of participants with observed data; SD, standard deviation; Q1, first quartile; Q3, third quartile.

**Supplemental Fig. 1 | Nausea and Use of AOMs on Days 1 and 30**

a) Use of anti-nausea medications, stratified by time on AOM (days 1 and 30)

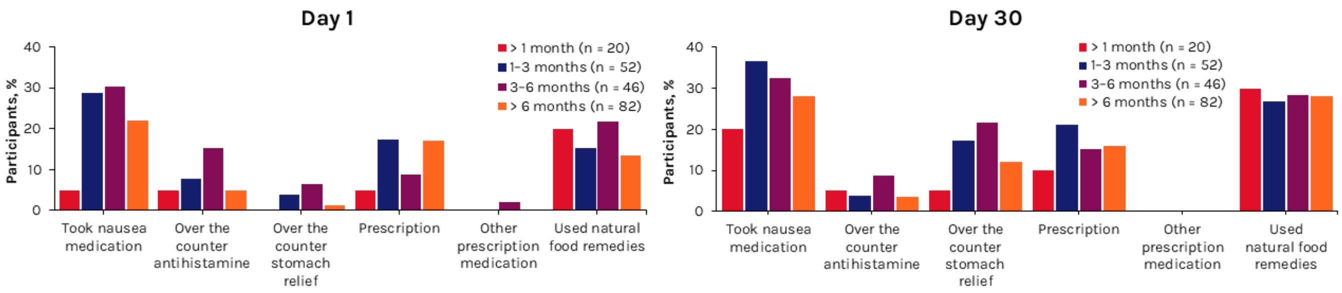


b) Days of nausea per month, stratified by time on AOM (days 1 and 30)

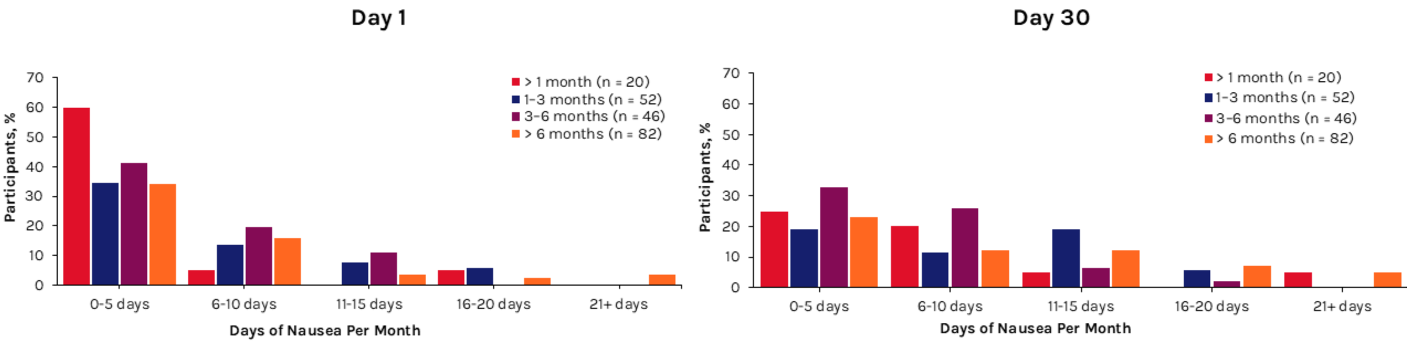


AOM, antiobesity medication.

**Supplemental Fig. 2 | Daily Nausea Diary Comparisons Based on AOM Duration**

a) Nausea severity,* by AOM duration

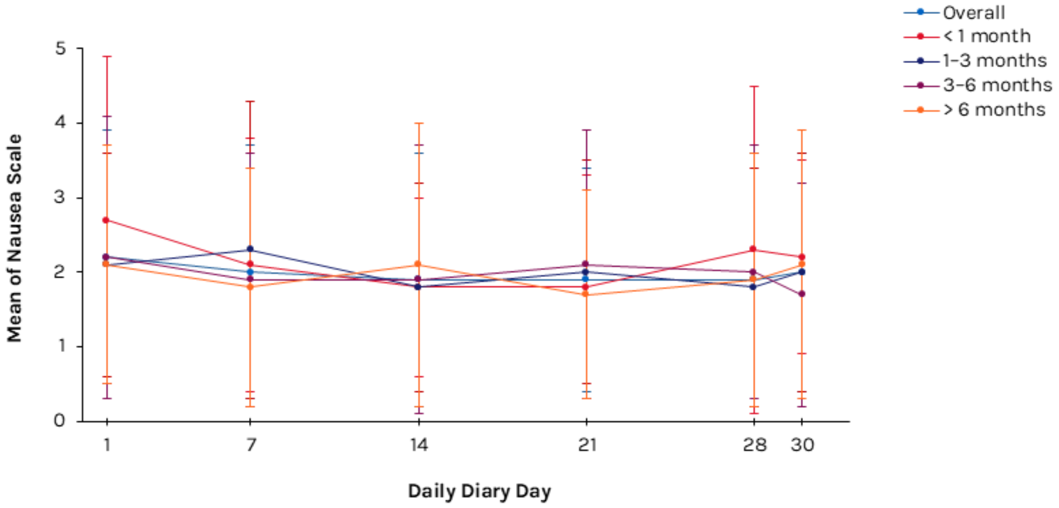


| | | | | | | |
|---|---|---|---|---|---|---|
| Overall, n = | 189 | 179 | 173 | 172 | 169 | 169 |
| < 1 month, n = | 18 | 17 | 17 | 17 | 16 | 16 |
| 1-3 months, n = | 49 | 44 | 43 | 43 | 41 | 41 |
| 3-6 months, n = | 44 | 43 | 42 | 42 | 42 | 42 |
| > 6 months, n = | 78 | 75 | 71 | 70 | 70 | 70 |

b) Negative impact of nausea, by AOM duration

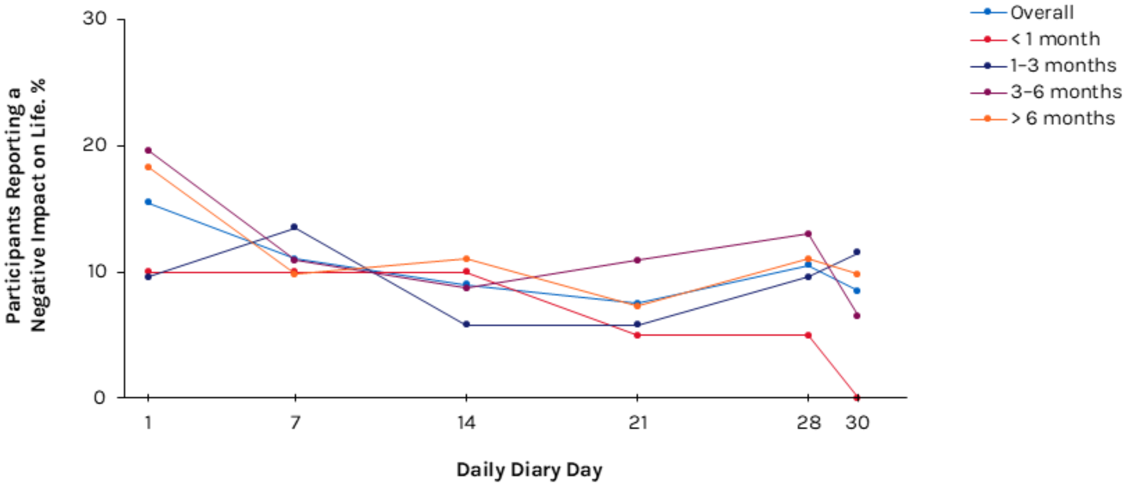


| | | | | | | |
|---|---|---|---|---|---|---|
| Overall, n = | 31 | 22 | 18 | 15 | 21 | 17 |
| < 1 month, n = | 2 | 2 | 2 | 1 | 1 | 0 |
| 1-3 months, n = | 5 | 7 | 3 | 3 | 5 | 6 |
| 3-6 months, n = | 9 | 5 | 4 | 5 | 6 | 3 |
| > 6 months, n = | 15 | 8 | 9 | 6 | 9 | 8 |

c) Use of anti-nausea medications, by AOM duration

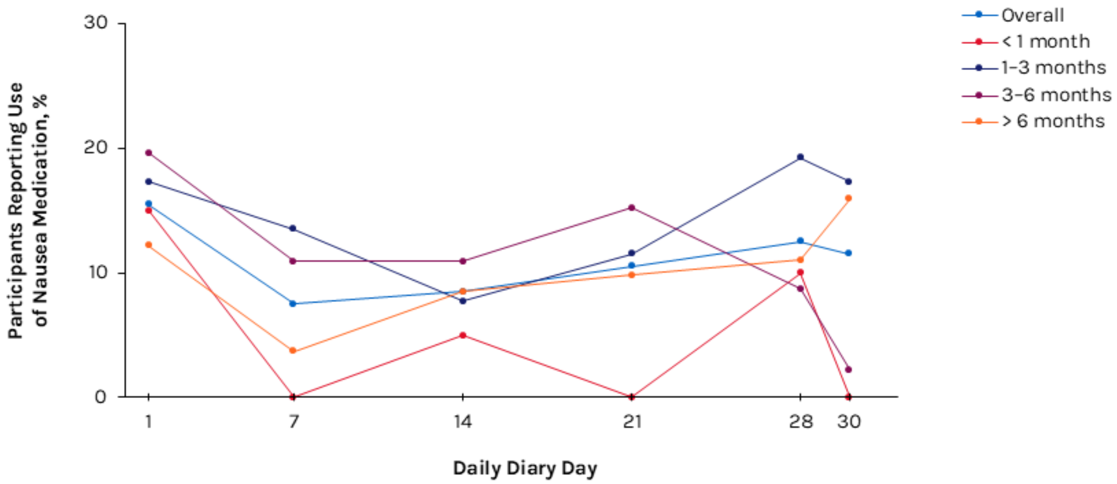


| | 1 | 7 | 14 | 21 | 28 | 30 |
|---|---|---|---|---|---|---|
| Overall, n = | 31 | 15 | 17 | 21 | 25 | 23 |
| < 1 month, n = | 3 | 0 | 1 | 0 | 2 | 0 |
| 1–3 months, n = | 9 | 7 | 4 | 6 | 10 | 9 |
| 3–6 months, n = | 9 | 5 | 5 | 7 | 4 | 1 |
| > 6 months, n = | 10 | 3 | 7 | 8 | 9 | 13 |

* Rated 1 to 10; 1 being the best, and 10 being the worst.
AOM, antiobesity medication.

### Supplemental Appendix 1 | Survey Questions

**Q1.** What is you height ? feet___________ inches ____________

What is your current Weight ? __________pounds

**Q2.** Which medication are you taking for weight loss?

A. Yes, Zepbound (tirzepatide)
B. Yes, Wegovy (semaglutide)

**Q3.** How long have you been on this medication?

A. Less than 1 month
B. 1 to 3 months
C. 3 to 6 months
D. More than 6 months

**Q4.** Are you taking this medication for weight management?

A. Yes, for weight management
B. Yes, for weight management and diabetes
C. No, just for diabetes
D. For another reason (please specify): ______________________

**Q5.** How motivated are you to lose weight? Please use a scale from 0 to 10, where 0 – Not motivated at all and 10 – Extremely motivated.

**Q6.** How long do you expect to take the medication you are on for weight loss?

A. 1 month
B. Up to 6 months
C. Up to a year
D. Up to two years
E. Until I've lost enough weight
F. I'm not sure yet

**Q7.** Do you plan to stop taking the weight loss medication after you achieve a certain weight?

A. Yes
B. No

**Q8.** If you were to use this medication long-term, what is the most ideal frequency for dosing?

A. Once a day
B. Once a week
C. Once a month
D. Once every 2 months
E. Once every 3 months (quarterly)
F. Once every 6 months (biannually)
G. Once a year (annually)

**SHOW TEXT:** For the next questions, think back over the last 30 days as best you can recall, and consider your nausea to answer these questions about how severe it was and the impact it has had on your life.

**Q9.** How severe was your nausea on a scale of 1 to 10: 1 = no nausea 5 = moderate nausea 10 = intolerable nausea

**Q10.** What did you do to improve your symptoms? (select all that apply)

A. Took nausea medication **(if selected, show next question)**
B. Used natural food remedies
C. Contacted my doctor
D. Stopped taking my anti-obesity medication (tirzepatide, semaglutide)
E. Delayed taking my anti-obesity medication (tirzepatide, semaglutide)
F. Complete skipped a dose of my anti-obesity medication (tirzepatide, semaglutide)
G. Nothing
H. I had no nausea

**Q11.** (Jump Logic: skip if previous answer did not include A)
Which anti-nausea medication did you use?

A. Over the counter antihistamine (diphenhydramine/Benadryl, meclizine, Dramamine)
B. Over the counter stomach relief (Emetrol, Pepto Bismol)
C. Prescription (ondansetron/Zofran, granisetron/Sustol, dolasetron/Anzemet, palonosetron/Aloxi)
D. Other prescription medication from my doctor/HCP

**Q12.** How many days would you estimate you felt nauseous?

A. 0 – 5 days
B. 6 – 10 days
C. 11 – 15 days
D. 16 – 20 days
E. 21+ days

**Q13.** How would you describe the nausea?

F. I had it the first few weeks, then it went away
G. I have it every time I take the medication
H. I have not had nausea
I. Other: ____________

**SHOW TEXT:** Consider your body weight or body shape to answer the following. Does your weight or body shape bother you in the areas listed below? questions. (Mark the options that best describes your current situation)

HCP, healthcare provider.

**Supplemental Appendix 2 | Nausea Scale**

**Show Text:** For the next 30 days, we'd like to ask you questions about your nausea symptoms and the effects it has on you. We will send you a daily form to complete which will ask you about your nausea symptoms for that day. The questions sent to you will be as per the below for each day

| Day | Nausea Score | Negative impact on life (physical activity, body pain, beahviro, sleep, sex, social interactions, work/school self-esteem) | Used nausea medication or food remedy | Skipped a dose |
|---|---|---|---|---|
| | 1 = none<br>10 = worst | Yes<br>No | Yes<br>No | Yes<br>No |
| 1 | | | | |

**Supplemental Appendix 3 | Patient-Reported Outcomes Survey**

| Patient Reported Outcomes in Obesity (PROS) | | | | | |
|---|---|---|---|---|---|
| 1 | Common physical activities (walking, climbing stairs and similar) | Considerably bothered | Moderately bothered | Mildly bothered | Not bothered |
| 2 | Bodily pain | Considerably bothered | Moderately bothered | Mildly bothered | Not bothered |
| 3 | Discrimination or discourteous behavior | Considerably bothered | Moderately bothered | Mildly bothered | Not bothered |
| 4 | Sleep | Considerably bothered | Moderately bothered | Mildly bothered | Not bothered |
| 5 | Sexual life | Considerably bothered | Moderately bothered | Mildly bothered | Not bothered |
| 6 | Normal social interaction | Considerably bothered | Moderately bothered | Mildly bothered | Not bothered |
| 7 | Work, school or other daily activities | Considerably bothered | Moderately bothered | Mildly bothered | Not bothered |
| 8 | Self-esteem | Considerably bothered | Moderately bothered | Mildly bothered | Not bothered |

**Supplemental Appendix 4 | Partners in Health Scale**

| Partners in Health scale: self-care questions (PiH) | | | | | | | | | | |
|---|---|---|---|---|---|---|---|---|---|---|
| 1 | Overall, what I know about my health condition(s) is: | 0 –0 – very little | 1 | 2 | 3 | 4 | 5 | 6 | 7 | 8 = a lot |
| 2 | Overall, what I know about the treatment, including medication of my health condition(s) is: | 0 – very little | 1 | 2 | 3 | 4 | 5 | 6 | 7 | 8 = a lot |
| 3 | I take medications or carry out the treatments asked by my doctor or health worker: | 0 – never | 1 | 2 | 3 | 4 | 5 | 6 | 7 | 8 = always |
| 4 | I share decisions made about my health condition(s) with my doctor or health worker: | 0 – never | 1 | 2 | 3 | 4 | 5 | 6 | 7 | 8 = always |
| 5 | I am able to deal with health professionals to get the services I need that fit with my culture, values, and beliefs: | 0 – not very well | 1 | 2 | 3 | 4 | 5 | 6 | 7 | 8 = very well |
| 6 | I attend appointments as asked by my doctor or health worker: | 0 – never | 1 | 2 | 3 | 4 | 5 | 6 | 7 | 8 = always |
| 7 | I keep track of my symptoms and early warning signs (blood sugar levels, peak flow, weight, shortness of breath, pain, sleep problems, mood): | 0 – never | 1 | 2 | 3 | 4 | 5 | 6 | 7 | 8 = always |
| 8 | I take action when my early warning signs and symptoms get worse: | 0 – never | 1 | 2 | 3 | 4 | 5 | 6 | 7 | 8 = always |
| 9 | I manage the effect of my health condition(s) on my physical activity (walking, household tasks): | 0 – not very well | 1 | 2 | 3 | 4 | 5 | 6 | 7 | 8 = very well |
| 10 | I manage the effect of my health conditions(s) on how I feel (that is, my emotions and spiritual wellbeing): | 0 – not very well | 1 | 2 | 3 | 4 | 5 | 6 | 7 | 8 = very well |
| 11 | I manage the effect of my health condition(s) on my social life (how I mix with other people) | 0 – not very well | 1 | 2 | 3 | 4 | 5 | 6 | 7 | 8 = very well |
| 12 | Overall, I manage to live a healthy life – no smoking, moderate alcohol, healthy food, regular physical activity, manage stress: | 0 – not very well | 1 | 2 | 3 | 4 | 5 | 6 | 7 | 8 = very well |

### Supplemental Appendix 5 | TAP® Survey Questions

6. On a scale from 0-10, how likely is it that you would recommend the TAP® device to a friend or colleague?

Not at all likely — Extremely likely

| 0 | 1 | 2 | 3 | 4 | 5 | 6 | 7 | 8 | 9 | 10 |
|---|---|---|---|---|---|---|---|---|---|---|

7. The TAP® device was easy to use by following the written Instructions for Use.

| Strongly disagree | Disagree | Neutral | Agree | Strongly agree |
|---|---|---|---|---|
| 1 | 2 | 3 | 4 | 5 |

TAP®, Touch Activated Phlebotomy.
